# Improving Model Transferability for Clinical Note Section Classification Models Using Continued Pretraining

**DOI:** 10.1101/2023.04.15.23288628

**Authors:** Weipeng Zhou, Meliha Yetisgen, Majid Afshar, Yanjun Gao, Guergana Savova, Timothy A. Miller

## Abstract

**Objective:** The classification of clinical note sections is a critical step before doing more fine-grained natural language processing tasks such as social determinants of health extraction and temporal information extraction. Often, clinical note section classification models that achieve high accuracy for one institution experience a large drop of accuracy when transferred to another institution. The objective of this study is to develop methods that classify clinical note sections under the SOAP (“Subjective”, “Object”, “Assessment” and “Plan”) framework with improved transferability.

**Materials and methods:** We trained the baseline models by fine-tuning BERT-based models, and enhanced their transferability with continued pretraining, including domain adaptive pretraining (DAPT) and task adaptive pretraining (TAPT). We added out-of-domain annotated samples during fine-tuning and observed model performance over a varying number of annotated sample size. Finally, we quantified the impact of continued pretraining in equivalence of the number of in-domain annotated samples added.

**Results:** We found continued pretraining improved models only when combined with in-domain annotated samples, improving the F1 score from 0.756 to 0.808, averaged across three datasets. This improvement was equivalent to adding 50.2 in-domain annotated samples.

**Discussion:** Although considered a straightforward task when performing in-domain, section classification is still a considerably difficult task when performing cross-domain, even using highly sophisticated neural network-based methods.

**Conclusion:** Continued pretraining improved model transferability for cross-domain clinical note section classification in the presence of a small amount of in-domain labeled samples.

## Introduction and background

Electronic Health Record (EHR) systems contain important clinical information in unstructured text, and natural language processing (NLP) is an important tool for its secondary use. Clinical note section classification is a foundational NLP task, as it facilitates many downstream tasks, and section information has been found beneficial for a diversity of clinical NLP tasks including named entity recognition [1], abbreviation resolution [2], cohort retrieval [3] and temporal relation extraction [4]. In this work we describe experiments on the task of clinical note section classification [5], using the SOAP (“Subjective”, “Objective”, “Assessment” and “Plan”) note framework to label note sections. In clinical practice, SOAP-style notes are widely used note-writing format taught for documenting the daily care of patients [6], [7]. Automatically classifying sections into SOAP categories is beneficial for better understanding the sourcing of information extracted by other NLP systems. For example, SDOH information may be more likely to be found in the social history section of a clinical note which is a “Subjective” section in the SOAP framework. Medication mentions may have different interpretation if they are in an “Objective” section (e.g., treatments in a medication list) versus a “Subjective” section (e.g., medication misuse in a social history). In addition, state-of-the-art NLP models (pre-trained transformers) have memory constraints that limit the number of words they can process [8], so processing only relevant sections may make these models more applicable.

Existing work in section classification [9], [10] has shown that the task is solvable for a given dataset, but that performance drops substantially when applying a trained system to a new dataset. In contrast to the SOAP task, those works used finer-grained section categories, which vary across datasets and have fewer training instances per label. In simplifying the section classification task to the SOAP classification task, we make it possible to perform more cross-domain experiments, and simplify the task to examine the cross-domain performance loss in a setting where we can eliminate one variable – the differences in the output space.

In clinical note section classification, researchers have found that statistical methods and modern pre-trained transformers (e.g., BERT [8]) achieved high performance for single institution modeling [9], [10]. In a study for classifying emergency departments reports into SOAP sections, researchers built a SVM classifier with lexical syntactic, semantic, contextual and heuristic features with SVM and the macro-F1 score was 0.85 [11]. In Rosenthal et al. [10], BERT achieved 0.99 and 0.9 F1 score for two section classification datasets with fine-grained section names. In Tepper et al. [9], researchers studied performing note segmentation and section classification together with fine-grained section names. Maximum Entropy Classifiers with fine-grained features (e.g., capital letters, numbers, blank lines, previous section names) achieved an F1 score of over 0.9 for two discharge summary datasets and one radiology report dataset. When transferring models learned from one dataset to another, the F1 score dropped to 0.6.

Domain adaptation refers to the study of improving model’s transferability from a source dataset to a target dataset and is a common theme in clinical NLP. In a study for psychiatric notes deidentification, three domain adaptation techniques, instance pruning, instance weighting, and feature augmentation were applied to a conditional random field (CRF) model for improving its adaption to the target dataset [12]. In Li et al. [13], researchers improved model adaption by training models on multiple domains and creating an ensemble. In Xing et al. [14], multi-task learning was applied on the task of segmenting words in Chinese medical text as a domain adaptation method. The model was trained on multiple tasks with the goal of learning the domain invariant features.

### Objective

The objective of this study is to develop methods that classify clinical note sections with SOAP (“Subjective”, “Object”, “Assessment” and “Plan”) labels. The secondary objective of this work is to examine the generalizability of existing datasets and methods by performing cross-domain validation, and to attempt to address any performance degradation with domain adaptation methods.

## Methods

### Datasets

We used three independent datasets across multiple health systems and different note types. The first dataset (**discharge**) consists of discharge summaries from the i2b2 2010 challenge from Partners Healthcare and Beth Israel Deaconess Medical Center [9]. The second dataset (**thyme)** includes colorectal clinical notes of the THYME (Temporal History of Your Medical Events) corpus of Mayo Clinic data [15]. The third dataset (**progress**) consists of MIMIC-III progress notes derived from providers across different specialty intensive care units [16], [17]. We created classification instances for each dataset by extracting sections from all the notes. While all three datasets had available section label annotations, the section labels were different across datasets. To facilitate cross-domain experiments, an expert physician informaticist (MA) mapped each dataset’s section labels into SOAP (“Subjective”, “Object”, “Assessment” and “Plan”) labels [11], [18]. The sections that did not fit into the SOAP framework (e.g., comments, addendum) were labeled as “Others”. This created a 5-way classification instance for each section. Table 1 presents the size, average word count, label distribution and train/test split ratio for each dataset. During SOAP mapping, we observed that some section headers covered both “Assessment” and “Plan” contents (e.g., the “Assessment and Plan” section label in the **progress** dataset). We mapped such sections to the “Assessment” label. As a result, the **progress** dataset has a section count of 0 for the “Plan” category in Table 1. When splitting the dataset into training and test set, for **discharge**, we randomly split the dataset with a 0.8/0.2 ratio. For **thyme** and **progress**, we followed the original train/test splits[15], [17].

**Table 1.** Size, average section word count, and label distribution of the **discharge, thyme** and **progress** dataset.

| dataset | total section counts | average word count | “Subjective” section count | “Objective” section count | “Assessment” section count | “Plan” section count | “Others” section count | train/ test split |
| --- | --- | --- | --- | --- | --- | --- | --- | --- |
| discharge | 1372 | 61 | 318 | 686 | 243 | 103 | 22 | 0.8/0.2 |
| thyme | 4223 | 74 | 1878 | 1329 | 676 | 100 | 240 | 0.73/0.27 |
| progress | 13367 | 46 | 4521 | 7039 | 787 | 0 | 1020 | 0.89/0.11 |

### In-domain and cross-domain section classification

We used the pre-trained transformer framework for section classification. We fine-tuned BioBERT [19] for the **thyme, discharge** and **progress** datasets. We used BioBERT as the BERT implementation because BioBERT was pretrained using biomedical texts and performed better than BERT on a variety of biomedical NLP tasks, including named entity recognition, relation extraction and question answering [19]. Other domain-appropriate BERT variants (e.g., BioClinicalBERT) are already pre-trained on MIMIC-III, the source of our **progress** dataset, so we avoid those models for the initial fine-tuning experiments to avoid data leakage.

We first measured the classification performance for the three datasets, both in the in-domain and cross-domain settings. These performance values represented the upper and lower bounds for our subsequent experiments. We measured the in-domain classification performance by testing the fine-tuned model on the same dataset’s test set. We measured the cross-domain classification performance by testing the fine-tuned model on the other two datasets’ test sets. We defined source domain as the dataset used for model fine-tuning, and target domain as the dataset used for model testing. We denote an experiment as FT_source_ if the model was fine-tuned on a source domain and tested on a target domain different from the source; we denote an experiment as FT_target_ if the model was both fine-tuned and tested on the same domain.

When fine-tuning BERT, we used a learning rate of 1e-5, epoch size of 40 and batch size of 10. These hyperparameters were tuned using the training set. The best model during model training (determined by the best F1 score on the held-out validation set) was saved and used for testing. The same hyperparameter settings are used in the cross-domain experiments to simulate the realistic case where target domain resources are usually too limited to conduct individualized hyperparameter search. The micro-F1 score (referred to as F1 score in future sections) was used as the evaluation metric. We implemented the Huggingface Transformers pipeline with AdamW optimizer for fine-tuning [20]. Experiments in this study were done on a 24GB NVIDIA TITAN RTX GPU with FP16 precision.

### Continued pretraining

Recent work has provided evidence that continued pretraining of pretrained language models on a target domain allows for better adaptability of the model [21]. Domain-adaptive pretraining (DAPT) is an unsupervised domain adaptation technique where a pre-trained model is trained for additional steps, using the same pre-training task of masked language modeling objective, on a large collection of unlabeled data from the target domain. Task-adaptive pretraining is similar, but uses a smaller amount of target domain data – only that portion that was labeled for the task of interest. For example, for the **progress** dataset, the domain-adaptive pretraining used the entire MIMIC-III dataset, and the task-adaptive pretraining considered the training set of **progress**. In previous work on general domain datasets [21], both DAPT and TAPT improved better cross-domain performance, and combining them sequentially (i.e. DAPT+TAPT) obtained the best performance. We thus experimented with pre-trained transformer models that have been adapted either with DAPT or DAPT+TAPT. In these experiments, the DAPT, TAPT, or DAPT+TAPT training is done on top of a base language model (BioBERT), followed by fine-tuning BERT on labeled examples in a source and/or target domain (as in the FT_source_ experiments in the last section).

We denote these experiments as DAPT + FT_source_ and DAPT + TAPT + FT_source_ in the remainder of the paper.

We note that existing work in the clinical domain could be interpreted as DAPT. For example, BioClinicalBERT[22] was created by doing continued pretraining on MIMIC-III [16] using BioBERT [19] as a starting point. From the perspective of downstream tasks that use MIMIC-III as a target domain (e.g., the **progress** dataset), comparing a BioBERT that has been fine-tuned on a source domain to BioClinicalBERT that has been fine-tuned on a source domain is essentially testing DAPT. Since BioClinicalBERT has already been shown to perform well on multiple tasks, in this work we use the existing BioClinicalBERT checkpoint as our DAPT model when **progress** is the target domain. When **thyme** is the target domain, we used an unreleased section of additional unlabeled notes for the patients in the THYME labeled corpus [15] to perform the continued pre-training for DAPT. For **discharge**, no additional unlabeled data is available. As a proxy, we again used MIMIC-III and used BioClinicalBERT as the DAPT model for **progress**.

In DAPT pretraining for **thyme**, we followed the setup of the BioClinicalBERT paper [22] and used a maximum training step count of 15000 and a learning rate of 5e-5. For TAPT, we followed the continued pretraining paper [21] and trained the model on the labeled data from the target domain (with the masked language modeling task, so it is still unsupervised) for 100 epochs with other settings being the same.

Our TAPT experiments used only the training splits of the **discharge, progress, and thyme** datasets.

To summarize our experimental settings, Table 2 presents the configuration details of experiments for when the **thyme** dataset is the target domain. The corresponding tables for **discharge** and **progress** datasets are included in Online Supplement.

**Table 2.**
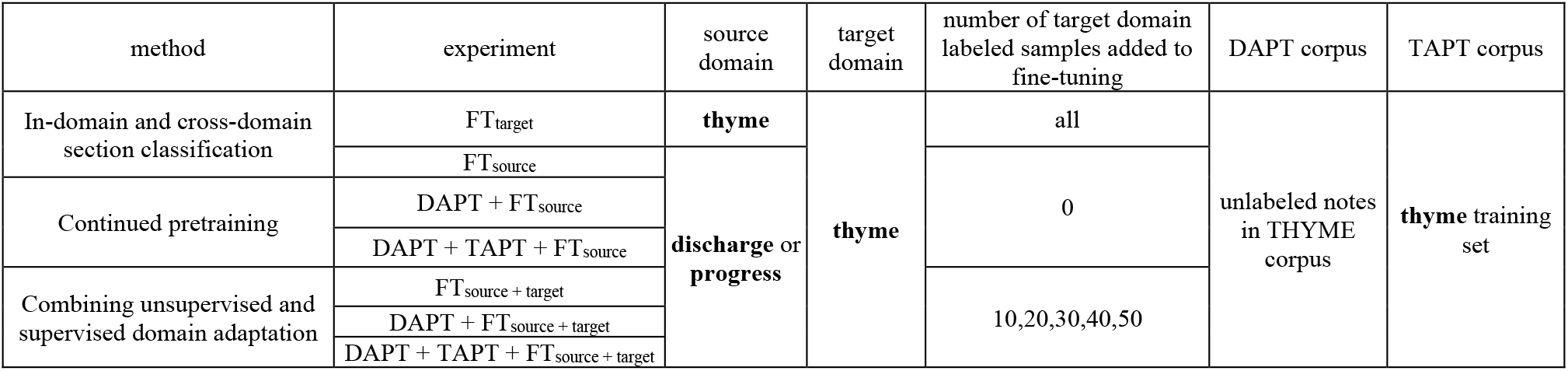
Description of in-domain and cross-domain experiments with **thyme** being the target domain.

### Combining unsupervised and supervised domain adaptation

In the DAPT and DAPT+TAPT experiments, we used only the source domain data for BERT fine-tuning, simulating the realistic setting where no annotation is possible at the target site (i.e., unsupervised domain adaptation). We next performed experiments that simulate the possibility that a small amount of labeled data is available at the target site, by including small numbers of labeled samples from the target domain during BERT fine-tuning (i.e., supervised domain adaptation). We also explore how the addition of labeled target domain data interacts with DAPT and TAPT. We varied the number of target domain samples from 10, 20, 30, 40 to 50. We denote these experiments as FT_source+target_, DAPT+FT_source+target_, and DAPT+TAPT+FT_source+target_.

### Quantifying the value of unsupervised domain adaptation

Both unsupervised and supervised domain adaptation are expected to provide performance increases over no adaptation, but they both require additional effort and have trade-offs in terms of implementation difficulty. If a practitioner is looking for guidance on whether to do continued pre-training or more data labeling, it would be useful to compare the value of these different methods in the same units. To facilitate this comparison, we analyzed our previous experiments to measure the value of unsupervised domain adaptation in terms of its equivalence to a number of labeled target domain samples. For example, if the FT_source+target_ model obtained an F1 score of 0.7 with 10 labeled target samples, and FT_target_ has an F1 score of 0.68 with 59 labeled samples, and 0.71 with 60 labeled samples, it means the value of the source domain training is equivalent to 60-10=50 additional target samples.

To calculate these values, we first extended our FT_target_ experiments on labeled data amounts ranging from 10 to 200 with an interval of 10, computing the F1 score for each experiment. We then linearly interpolate between consecutive labeled data amounts (e.g., between 10 and 20), which allows us to create a function *f1*_*target*_(*n*) that returns an estimated F1 score for every whole number *n* of labeled data amounts between 10 and 200. While this function is not invertible, we can create a pseudo-inverse:

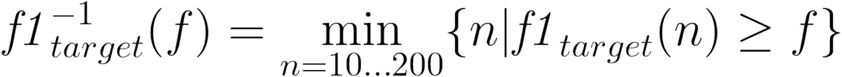

which, given an F1 score *f*, returns the lowest number of labeled target instances in the FT_target_ experiment that matched or exceeded that score. Then, for each of the cross-domain settings (FT_source+target_, DAPT+FT_source+target_, and DAPT+TAPT+FT_source+target_), we have F1 scores for a range of 10 to 50 labeled data points from the experiments above. For each cross-domain setting and the corresponding target domain samples included (e.g., FT_source+target_, 10), we can get from *f1*^*-1*^_*target*_(f) the minimum number of target domain samples FT_target_ would need to match that score. For each setting we report the added value of the added component (e.g., for DAPT+FT_source+target_ we report the added value over FT_source+target_) to isolate the value of each intervention.

## Results

### In-domain and cross-domain section classification

Table 3 shows the results of the in-domain and cross-domain experiments with fine-tuning, DAPT+FT, and DAPT+TAPT+FT. When moving from in-domain to cross-domain, the F1 scores dropped from 0.97-0.99 range to 0.541-0.717 range. The average in-domain (FT_target_) F1 score is 0.977. The average cross-domain (FT_source_) F1 score is 0.618.

**Table 3.** F1 scores of in-domain and cross-domain models, with DAPT and TAPT when applicable. The best F1 score for each combination of source and target domain is in bold.

| source domain (→) | discharge |  |  | thyme |  |  | progress |  |  |
| --- | --- | --- | --- | --- | --- | --- | --- | --- | --- |
| target domain (↓) | FT | DAPT + FT | DAPT + TAPT + FT | FT | DAPT + FT | DAPT + TAPT + FT | FT | DAPT + FT | DAPT + TAPT + FT |
| discharge | 0.972 | - | - | 0.572 | 0.6 | **0.675** | **0.541** | 0.5 | 0.501 |
| thyme | **0.601** | 0.469 | 0.53 | 0.99 | - | - | **0.646** | 0.632 | 0.544 |
| progress | 0.656 | 0.67 | **0.749** | **0.717** | 0.58 | 0.528 | 0.973 | - | - |

### Continued pretraining

Table 3 also shows that continued pretraining led to a decreased performance when **thyme** was the target domain. The effect of continued pretraining was mixed for **progress** and **discharge**. No significant performance improvement was observed when continued pretraining (DAPT or DAPT+TAPT) was applied directly on cross-domain section classification.

### Continued pretraining and fine-tuning with target domain labeled data

Figure 1 shows learning curves when some target-domain labeled data was provided for fine tuning. When comparing before and after continued pretraining (DAPT or DAPT+TAPT), we found continued pretraining generally improved model performance when combined with small numbers of target domain instances.

**Figure 1.**
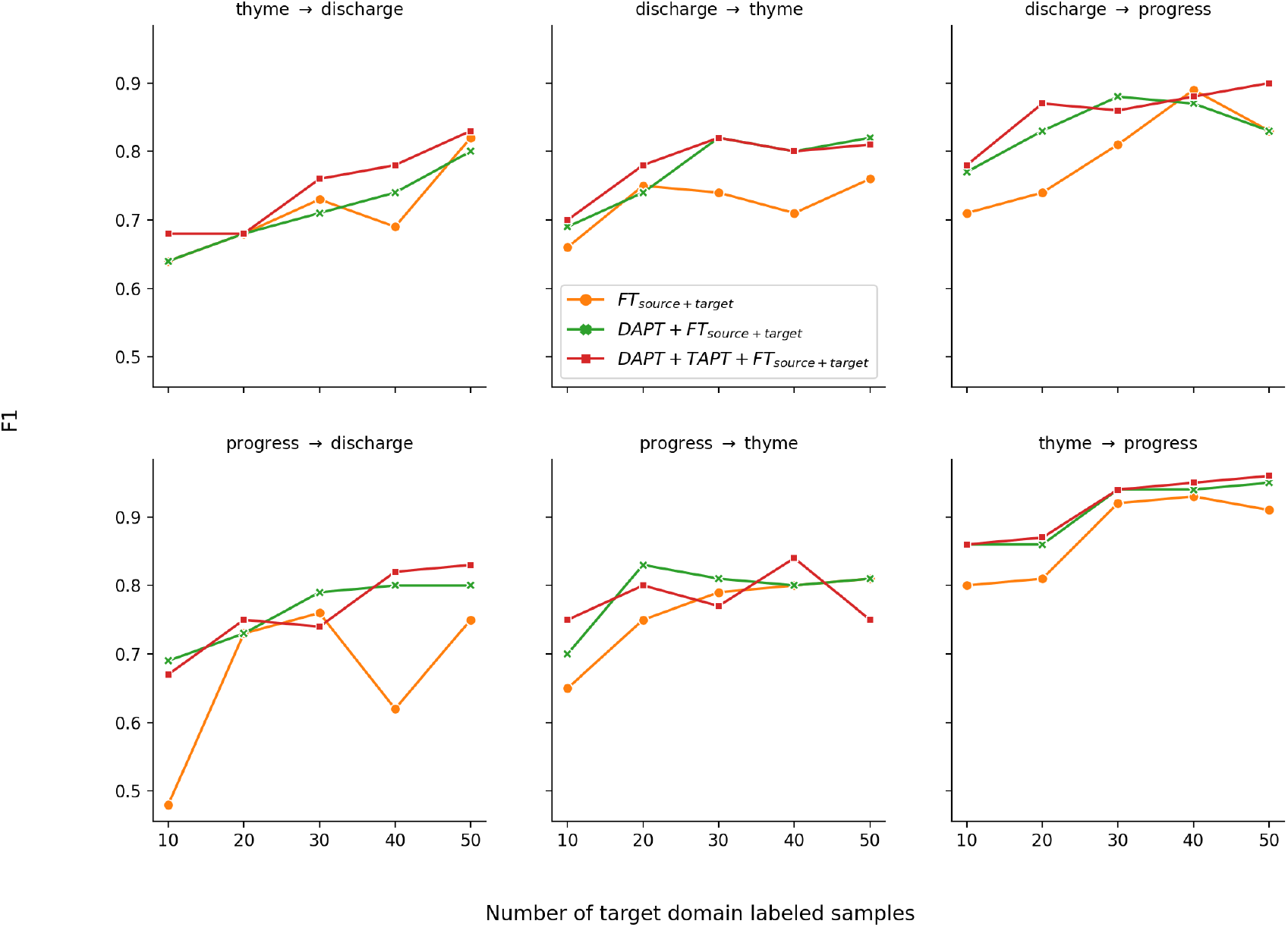
F1 scores of FT_source + target_, DAPT+FT_source + target_, and DAPT+TAPT+FT_source + target_ with 10, 20, 30, 40 and 50 target domain samples for different source and target domain experiments. For example, **thyme** → **discharge** represents the experiment with **thyme** being the source domain and **discharge** being the target domain.

Figure 2 shows the average learning curve across the 6 comparisons. On average, continued pretraining (DAPT or DAPT+TAPT) improved over the model without it (FT_source + target_) consistently. When comparing within continued pretraining models (DAPT FT_source + target_ and DAPT+TAPT+FT_source + target_), we found applying TAPT after DAPT further increased the F1 score for four out of five sample sizes.

**Figure 2.**
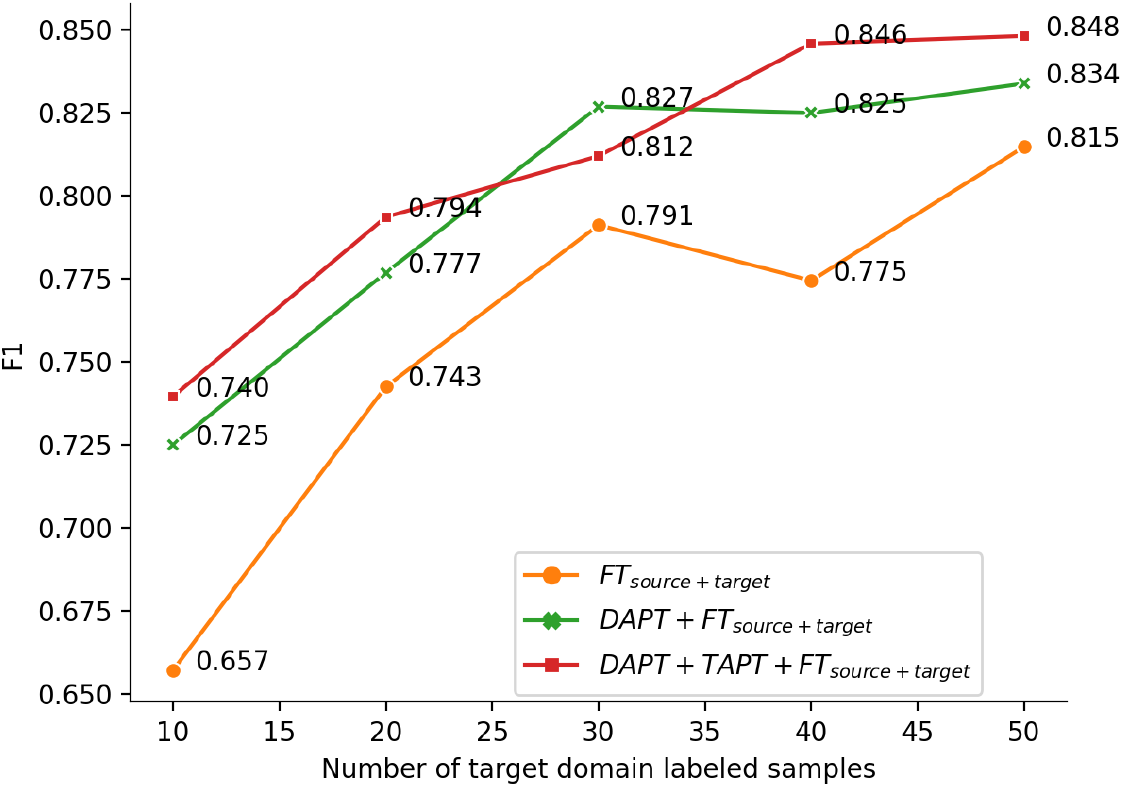
Dataset averaged F1 scores of FT_source + target_, DAPT+FT_source + target_, and DAPT+TAPT+FT_source + target_ with 10, 20, 30, 40 and 50 target domain samples included in fine-tuning.

### Quantifying the value of continued pretraining

Figure 3 visualizes the method for estimating the value of continued pre-training and the results from the DAPT+TAPT+FT_source+target_ experiment. First, the results from Figure 2 are overlaid with F1 scores from the FT_target_ experiments extended to use up to 200 target domain samples. We project horizontal lines from several points on the DAPT+TAPT+FT_source+target_ curve until they intersect with the FT_target_ curve. For example, at the left of the figure, DAPT+TAPT+FT_source+target_ achieved an F1 score of 0.74 when 10 target domain samples were included, and it intersects the FT_target_ curve when n=99. This corresponds to f1^-1^target(0.74) = 99, meaning the value of the transfer learning and pre-training is equivalent to an additional 89 target domain samples for FT_target_. The equivalent visualizations of the FT_source+target_, and DAPT+TAPT+FT_source+target_ curves are included in Online Supplement.

**Figure 3.**
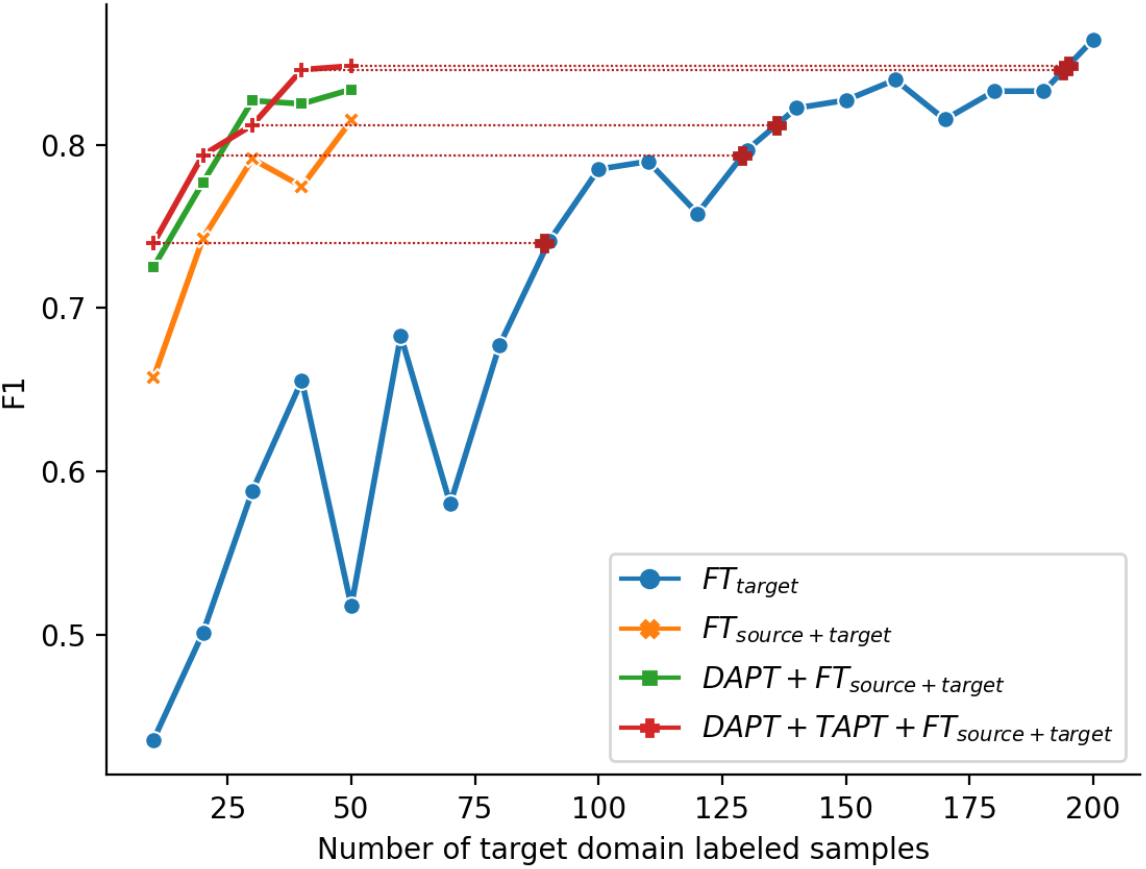
Dataset averaged F1 scores of FT_target_ with target domain labeled samples varying from 10 to 200, overlaying with Figure 2. Horizontal dotted lines between DAPT+TAPT+FT_souce+target_ and FT_target_ curves visualize applying f1^-1^target(f) on the F1 scores of DAPT+TAPT+FT_souce+target_ for obtaining the equivalent FT_target_ training sample size.

**Figure 4.**
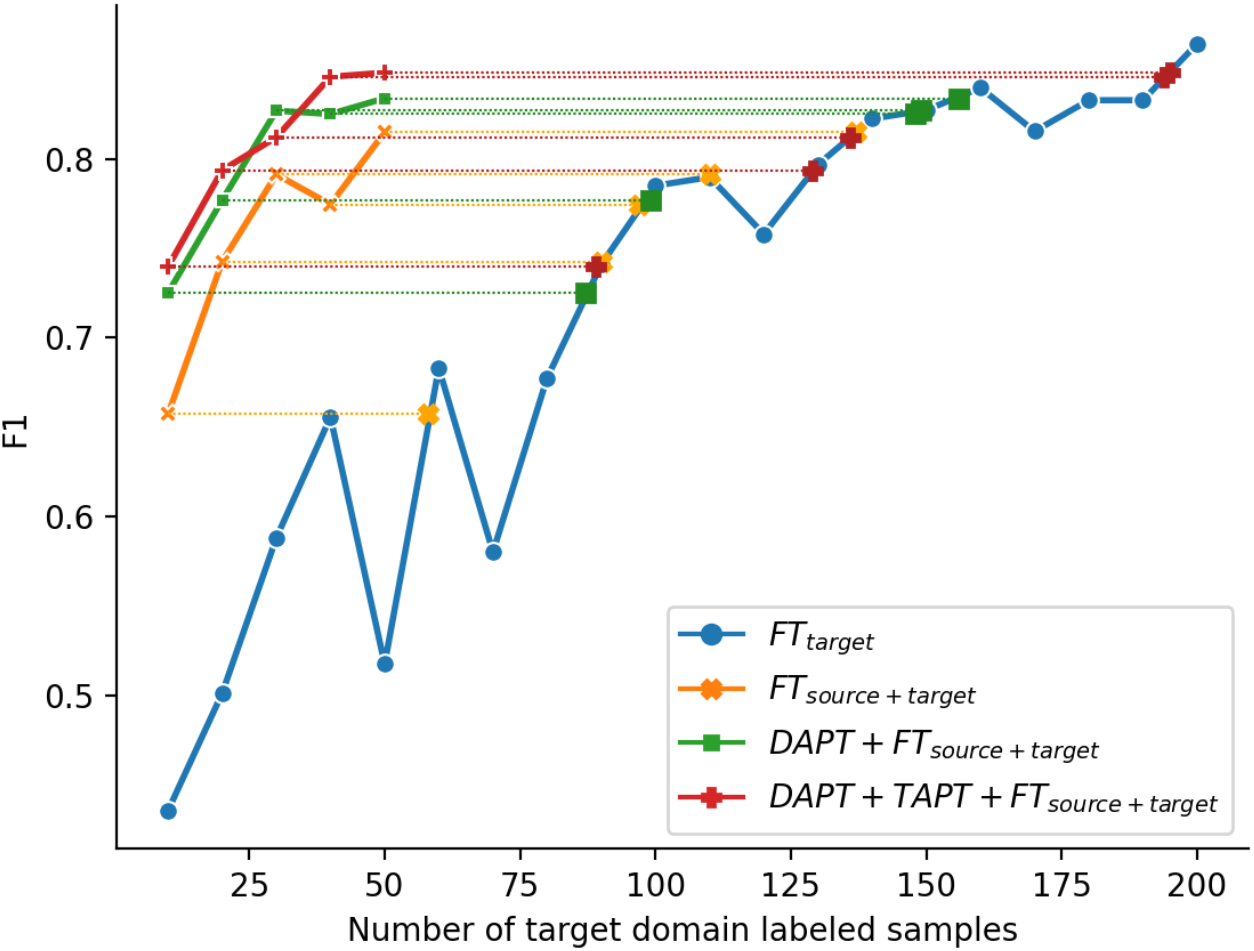
Dataset averaged F1 scores of FT_target_ with target domain labeled samples varying from 10 to 200, overlaying with Figure 2. Horizontal dotted lines between FT_souce+target_, DAPT +FT_souce+target_, and DAPT+TAPT+FT_souce+target_ and FT_target_ curves visualize applying f1^-1^target(f) on the F1 scores of FT_souce+target_, DAPT +FT_souce+target_, and DAPT+TAPT+FT_souce+target_ for obtaining the equivalent FT_target_ training sample size.

Table 4 shows the equivalent target domain sample size of the three cross-domain models, estimated by applying *f1*^*-1*^_*target*_*(f)* to every cross-domain setting with target domain sample size varying from 10 to 50 (and corresponding to the length of the horizontal lines in Figure 3). We averaged them over sample size, and by subtracting between incrementally different settings, we find 29.4 target domain samples being the added value of DAPT over FT_source+target_, and 50.2 being the added value of DAPT+TAPT over FT_source+target_.

**Table 4.** The effective target domain sample size of FT_source+target_, DAPT+FT_source+target_, and DAPT+TAPT+FT_source+target_ with target domain sample size varying from 10 to 50. The added value of DAPT and DAPT+TAPT over FT_source + target_ are shown in parenthesis.

| training strategy (→)<br>target domain (↓)<br>labeled sample size | $FT_{\text{source+target}}$ | $DAPT+FT_{\text{source+target}}$ | $DAPT+TAPT+FT_{\text{source+target}}$ |
| --- | --- | --- | --- |
| 10 | 58 | 87 | 89 |
| 20 | 90 | 99 | 129 |
| 30 | 110 | 149 | 136 |
| 40 | 97 | 148 | 194 |
| 50 | 137 | 156 | 195 |
| average | 98.4 | 127.8 (29.4) | 148.6 (50.2) |

## Discussion

Our results show that, while SOAP section classification is a straightforward task for humans, and one that can be effectively solved for individual datasets, current state of the art methods did not solve the task in a generalizable way. Part of the challenge may be attributable to different institutions having different documentation practices by providers, different note types in the EHR, and changes in label distribution. Many tasks are not adequately tested in out-of-sample environments across different domains and we provided a rigorous approach across multiple centers and note types to show that even “simple” tasks are difficult to generalize. The results also follow a similar finding in a finer-grained version of the task [9], as well as other clinical NLP tasks [23], but is perhaps more surprising here due to the relative simplicity of the task and the degree to which it is solved within each dataset. The attempts to leverage large language models and multiple fine-tuning and continual training approaches still did not completely overcome the cross-domain challenges.

The experiments between different combinations of training sets and training methods highlight trade-offs between different ways of mitigating the performance drop-offs when crossing domains. Unsupervised adaptation methods like DAPT and TAPT show benefits that are equivalent to dozens of target-domain training samples, but only when some target samples are already annotated. We also noted minimal performance gain from TAPT over DAPT, unlike prior work [21]. The small benefit from TAPT could be due to the fact that transfer learning already brought knowledge to the model in a similar form as pretraining. One important direction moving forward is to regularly report quantification of this type of information across tasks, so that different NLP tasks can be situated amongst each other in terms of the relative benefit they receive from unsupervised adaptation versus labeling additional instances.

The high value of unsupervised domain adaptation of pre-trained transformers is an encouraging result of this work. We caution, however, that it does not tell a complete story. Target domain annotation and continued pretraining, our two adaptation methods, both can be challenging and require resources at a target site. So, while the improvements of DAPT and TAPT are large in some cases, for this task they do seem to require some small amount of target-domain labeling. It could be the case that annotating a few hundred more instances is actually a more efficient decision than setting up continued pretraining infrastructure. In summary, even for the straightforward SOAP section classification task, these questions around adapting NLP systems are complex.

Each of the individual datasets we used were derived from single centers, which may be a contributing factor to the lack of generalizability. Future work in this task should explore the benefits of incorporating more variability in the types of notes and health systems used as source training data, to see whether combinations of datasets generalize better.

Future work should also extend to the segmentation version of the task, to see whether the same conclusions apply in that setting. Finally, future work should study whether the same findings may also be applicable to the more fine-grained section classification task, where the problem is more challenging due to lack of label standardization and sparsity of different section labels.

## Conclusion

The classification of clinical note sections is a critical step for downstream natural language processing tasks such as named entity recognition, cohort selection and temporal information extraction. In this study, we used continued pretraining to improve the transferability of such models. We studied three datasets from different institutions and found the average F1 score dropped from 0.977 to 0.618 when switching from in-domain to cross-domain prediction. We found that continued pretraining was not suitable when only source domain labeled samples were included in model training. When target domain labeled samples were included in model training, continued pretraining had an improvement on model transferability.

## Data Availability

All data produced in the present study are available upon reasonable request to the authors

## Acknowledgement

Research reported in this publication was supported by the National Library Of Medicine of the National Institutes of Health under Award Number R01LM012973, R01LM012918, and R01LM013486. The content is solely the responsibility of the authors and does not necessarily represent the official views of the National Institutes of Health.

## Online Supplement

**Table 5.**
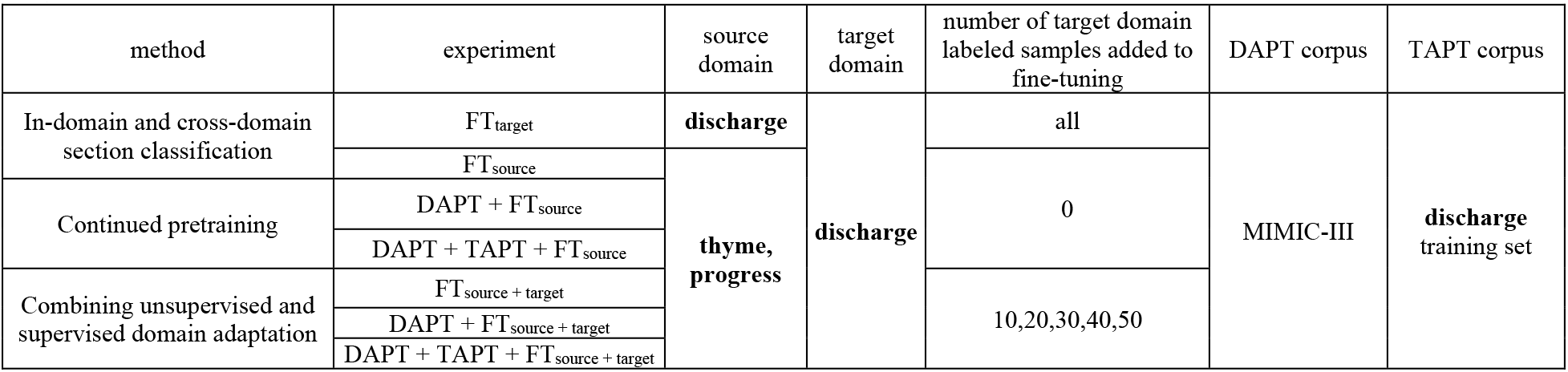
Description of in-domain and cross-domain experiments with **discharge** being the target domain.

**Table 6.**
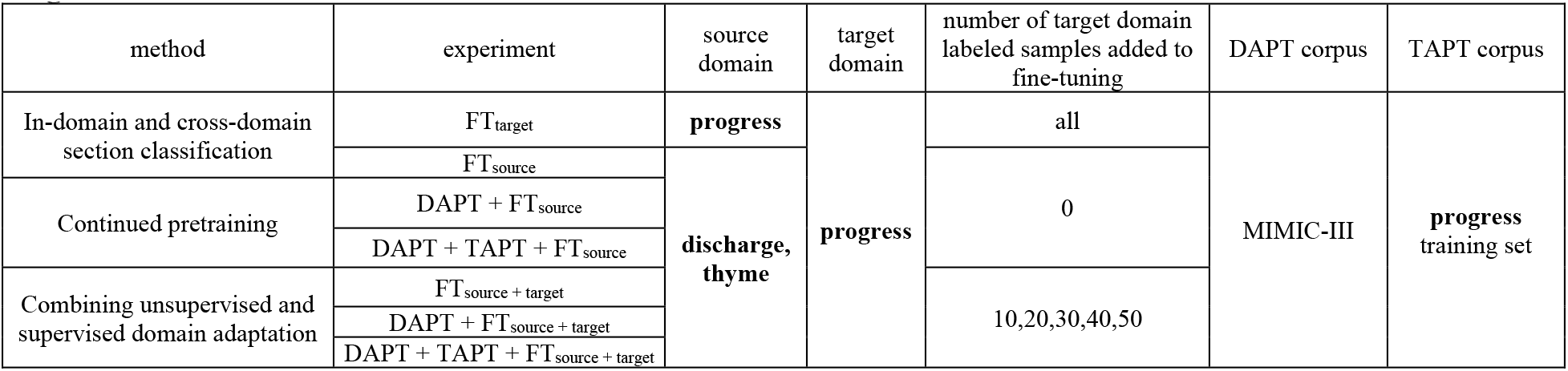
Description of in-domain and cross-domain experiments with **progress** being the target domain.

